# How do clinical researchers generate data-driven scientific hypotheses? Cognitive events using think-aloud protocol

**DOI:** 10.1101/2023.10.31.23297860

**Authors:** Xia Jing, Brooke N. Draghi, Mytchell A. Ernst, Vimla L. Patel, James J. Cimino, Jay H. Shubrook, Yuchun Zhou, Chang Liu, Sonsoles De Lacalle

## Abstract

**Objectives:** This study aims to identify the cognitive events related to information use (e.g., “Analyze data”, “Seek connection”) during hypothesis generation among clinical researchers. Specifically, we describe hypothesis generation using cognitive event counts and compare them between groups.

**Methods:** The participants used the same datasets, followed the same scripts, used VIADS (a <u>v</u>isual interactive <u>a</u>nalysis tool for filtering and summarizing large <u>d</u>ata <u>s</u>ets coded with hierarchical terminologies) or other analytical tools (as control) to analyze the datasets, and came up with hypotheses while following the think-aloud protocol. Their screen activities and audio were recorded and then transcribed and coded for cognitive events.

**Results:** The VIADS group exhibited the lowest mean number of cognitive events per hypothesis and the smallest standard deviation. The experienced clinical researchers had approximately 10% more valid hypotheses than the inexperienced group. The VIADS users among the inexperienced clinical researchers exhibit a similar trend as the experienced clinical researchers in terms of the number of cognitive events and their respective percentages out of all the cognitive events. The highest percentages of cognitive events in hypothesis generation were “Using analysis results” (30%) and “Seeking connections” (23%).

**Conclusion:** VIADS helped inexperienced clinical researchers use fewer cognitive events to generate hypotheses than the control group. This suggests that VIADS may guide participants to be more structured during hypothesis generation compared with the control group. The results provide evidence to explain the shorter average time needed by the VIADS group in generating each hypothesis.

**What is already known on this topic:** how hypotheses were generated when solving a puzzle or a medical case and the reasoning differences between experienced and inexperienced physicians.

**What this study adds:** Our study facilitates our understanding of how clinical researchers generate hypotheses with secondary data analytical tools and datasets, the cognitive events used during hypothesis generation in an open discovery context.

**How this study might affect research, practice, or policy:** Our work suggests secondary data analytical tools and visualization may facilitate hypothesis generation among inexperienced clinical researchers regarding the number of hypotheses, average time, and the cognitive events needed per hypothesis.

## Introduction

A research hypothesis is an educated guess regarding relationships among different variables [1,2]. A research question typically comprises one to several scientific hypotheses that drive the direction of most research projects [1,3-5]. If we consider the life cycle of a research project, hypothesis generation constitutes its starting point. Without a significant, insightful, and novel hypothesis to begin with, it is difficult to have an impactful research project regardless of the study design, experiment implementation, and results analysis. Therefore, hypothesis generation plays a critical role in a research project. There are several studies investigating the mechanism of the generation of scientific hypotheses by researchers, both in science (e.g., Dunbar and Khar [6,7]) and in clinical medicine (e.g., Joseph and Patel [8,9]). However, none of these studies address how an analytic tool can be used to facilitate the hypothesis-generation process.

At least two categories of hypothesis are used frequently in scientific research. One is a hypothesis originating from experimental observations, e.g., any unusual phenomena observed during experiments in the context of “wet lab”. The other category is a hypothesis originating from the context of data analysis, for example, studies in epidemiology, genomics, and informatics [10-12]. Observations of unique or unusual phenomena in the first category and observations of trends in the second category are both critical in developing hypotheses [7,13]. Herein, we focus on the hypothesis generation within the second category.

In the past decades, there has been much work toward understanding scientific thinking and reasoning, medical reasoning, analogy, and working memory [7,14]. Educational settings and math problems were used to explore the reasoning process [15-17]. However, scientific hypothesis generation was not addressed, and the mechanism of explicit cognitive processes during scientific hypothesis generation remains unclear. The main differences between scientific reasoning and hypothesis generation include: a) the starting points of the two processes are different; many studies involving scientific reasoning start from an existing problem or puzzle [17-20], whereas data-driven hypothesis generation searches for a problem or a focus area to begin, named as open discovery by; Henry et al. [21]; b) the mechanisms between the start and end points of the two processes may differ, with convergent thinking used more in scientific reasoning when a question or a puzzle needs to be solved [7] and divergent thinking used more in data-driven scientific hypothesis generation. Meanwhile, hypothesis generation in medical diagnosis starts with a presented medical case or symptoms [19,22], which is similar to scientific reasoning.

We previously developed a conceptual framework for scientific hypothesis generation and its contributing factors [23]. Researchers have explored the possibilities of automatically generating scientific hypotheses in the past [10,24-28]; however, these authors recognized the challenges faced by an automated tool for such an advanced cognitive process [24,29,30].

Our study aims to obtain a better understanding regarding the scientific hypothesis generation process in clinical research. Considering hypotheses can directly impact and guide the direction of any research project, the findings of this work can potentially impact the clinical research enterprise. The research protocol [31], VIADS [32-34] (a <u>v</u>isual interactive <u>a</u>nalytic tool for filtering and summarizing large health <u>d</u>ata <u>s</u>ets coded with hierarchical terminologies—VIADS, a secondary data analytical tool developed by our team) usability [35], and quality evaluation of the hypotheses generated by participants [23] have all been published. This manuscript focuses on the cognitive events used by experienced and inexperienced clinical researchers during hypothesis generation.

## Methods

### Study flow and data sets used

The 2 × 2 study compared the hypothesis generation process of the clinical researchers with and without VIADS on the same datasets (Appendix A), with the same study scripts (Appendix B), and within the same timeframe (2 hours/study session), and they all followed the think-aloud method. The participants were separated into experienced and inexperienced clinical researchers based on predetermined criteria[31], e.g., years of experience and number of publications as significant contributors. The data were extracted from the National Ambulatory Medical Care Survey (NAMCS) conducted by the Centers for Disease Control and Prevention in 2005 and 2015 [36]. We preprocessed the NAMCS data sets by calculating and aggregating the ICD-9-CM diagnostic and procedural codes and their frequencies. The participants were asked to analyze the data and generate hypotheses and articulate their mind and actions during the process, i.e., study sessions. The screen activities and conversations between participants and the study facilitator were recorded via BBFlashback. The recordings were transcribed by a professional service.

#### Cognitive events coding for the hypothesis generation recordings

Based on the experience of conducting all study sessions, initial data analysis, the feedback from the investigation team, and literature review [1,13,37-41], a preliminary conceptual framework of the cognitive hypothesis generation process was developed ***before*** coding (Figure 1). The conceptual framework served as a foundational framework to formulate the initial codes and code groups (Appendix C) that were used to code the transcriptions of the recordings, mainly for cognitive events (e.g., seek connections, analogy) in the hypothesis generation process. For example, “Analogy” was used when a participant compared one’s last study with the analysis results in front of him/her. “Use PICOT” was used when a participant used PICOT (i.e., patient, intervention, comparison, outcome, type of study) to formulate an idea into a formal hypothesis.

**Figure 1.**
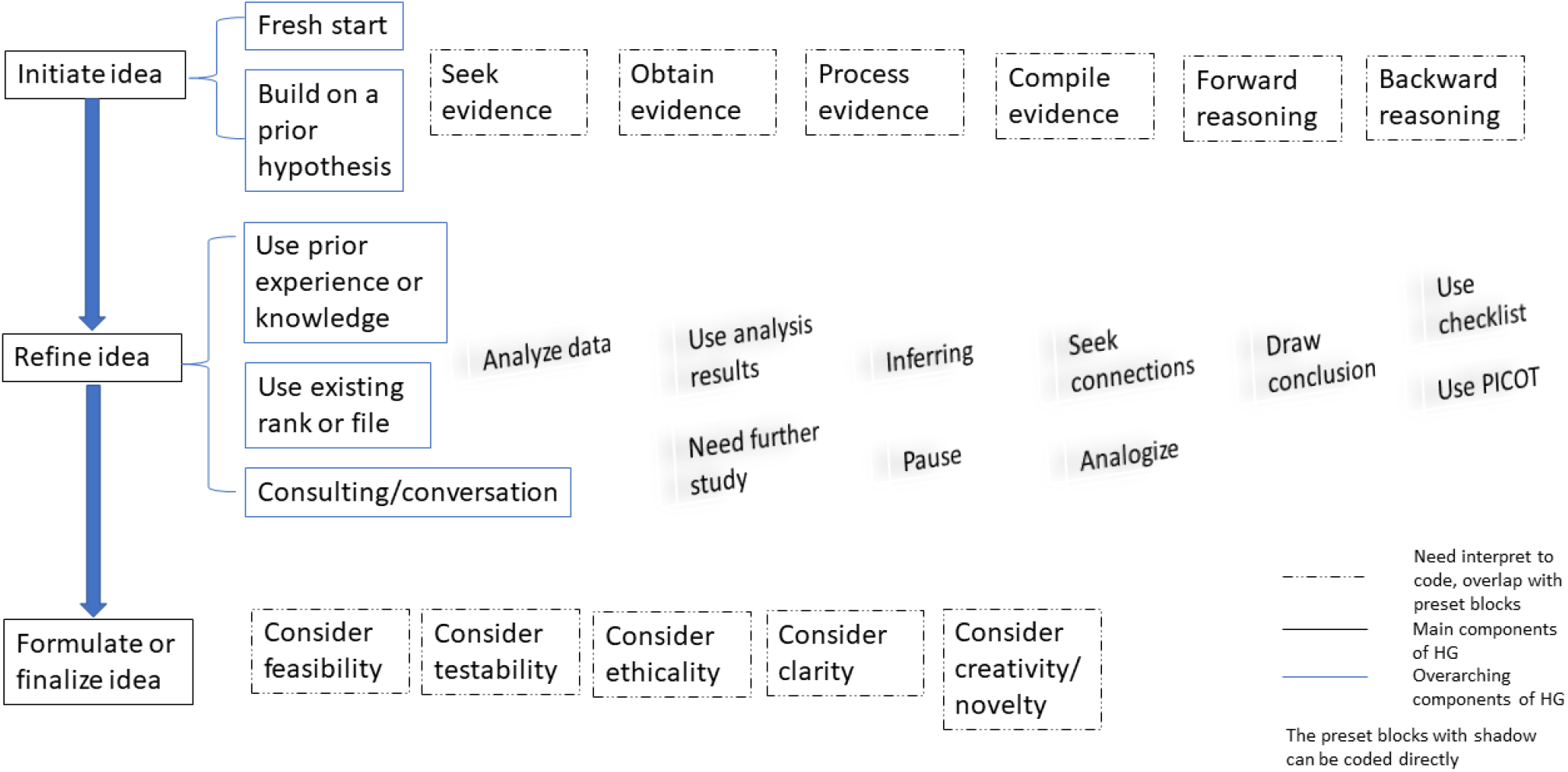
Initial version of the framework on cognitive events during hypothesis generation

The transcription of one study session was utilized as a pilot coding case to set the initial coding principles (Appendix D). The pilot coding sessions were used as training sessions for the two coders as well. The rest of the transcriptions were coded by the two coders independently and separately first. The two coders compared their coding results, discussed any discrepancies, and reached a consensus on coding later by including the study facilitator and modifying the coding principles.

More codes and code groups were added while the coding progressed. After coding all the study session transcripts, the two coders also organized each hypothesis generation as an independent process and labeled the cognitive events during each hypothesis generation. We investigated the possible hypothesis generation processes based on coded cognitive events.

#### Data analytics strategy

This study used the cognitive events and the aggregated frequencies of these events to demonstrate the possible hypothesis generation process. While analyzing the cognitive events, we considered the results from four levels: (1) each hypothesis generation as a unit and we examined all hypotheses (n = 199), (2) each participant as a unit and all participants (n = 16) as a unit, (3) the group of participants who used VIADS as a unit (n = 9), and (4) the group of participants who did not use VIADS as a unit (n = 7). Correspondingly, the results were also organized at these four levels. We performed independent t-tests to compare the cognitive events between participants (a) in the VIADS and control groups and (b) between the experienced (3 participants, 36 hypotheses) and inexperienced clinical researchers (13 participants, 163 hypotheses). The study sessions of two participants’ (in the control group, both were inexperienced clinical researchers) were missing from the coding data because of technical failures resulting in partial recording of their study sessions, and their data were excluded from the analysis.

All hypotheses were rated by an expert panel of seven members using the same metrics for quality evaluation [23,42]. We deemed a hypothesis as invalid if three or more experts rated it as 1 (the lowest rating) on validity (significance and feasibility are two additional dimensions used for evaluation) of the hypothesis. However, we ***included*** the analysis of the result for all the hypotheses and valid hypotheses.

### Ethics statement

The study was approved by the Institutional Review Board of Clemson University, South Carolina (IRB2020-056) and Ohio University Institutional Review Boards (18-X-192).

## Results

### Hypothesis generation framework

Figure 2 is a refined and evolving version of the initial framework shown in Figure 1, our preliminary understanding of hypothesis generation. Figure 2 was instrumental in directly guiding the coding of the cognitive events. The predominant cognitive events within the processing evidence category include “Using analysis results” (30%), “Seeking connections” (23%), and “Analyze data” (20.81%, Figure 2). Appendix E illustrates the processes and events used percentages while generating hypotheses. Appendix F presents individual cognitive events used for all hypotheses and valid hypotheses, respectively.

**Figure 2.**
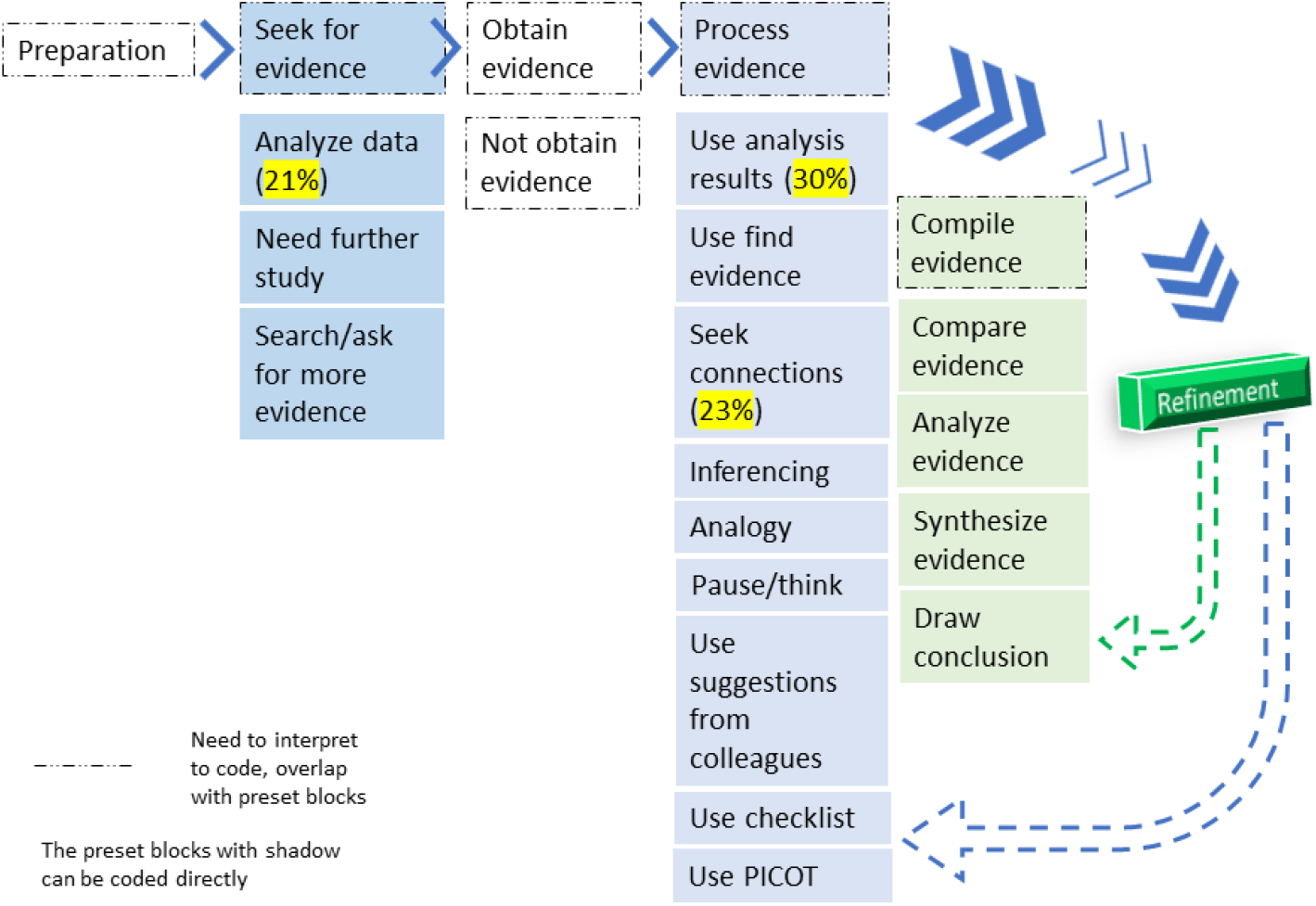
Cognitive process frameworks for scientific hypothesis generation in clinical research; the highest percentages of cognitive events used by clinical researchers were highlighted.

### Overall cognitive events usage during hypothesis generation

Sixteen participants generated 199 hypotheses during the 2-hour study sessions, with 163 originating from the inexperienced groups (Table 1). We used 20 distinct codes, i.e., cognitive events and 6 code groups (Figure 2). Appendix C showcases the comprehensive codebook. Appendix D delineates the rationale and principles established during the coding phase. In total, 1216 times of cognitive events were applied across the 199 hypotheses. On average, inexperienced clinical researchers in the control group applied 7.38 cognitive events per hypothesis. Conversely, inexperienced clinical researchers in the VIADS group used 4.48 (p< 0.001 versus control) cognitive events per hypothesis with the lowest standard deviation (SD, 2.43). Experienced clinical researchers employed 6.15 (p < 0.01 versus junior VIADS) cognitive events per hypothesis. Notably, the inexperienced clinical researchers in the control group demonstrated the highest average number of cognitive event usage with the largest SD (5.02), whether we considered all hypotheses or just valid ones (Table 1). The experienced participants have approximately 10% higher valid hypotheses (72.22% vs. 63.19%) than junior participants.

**Table 1.** Group-wise comparison of cognitive events used while generating hypotheses.

|  | All hypotheses | Valid # (%) | Invalid # (%) |
| --- | --- | --- | --- |
| **# Hypotheses by different groups** |  |  |  |
| # Hypotheses by all participants | 199 | 129 (64.82) | 70 (35.18) |
| # Hypotheses by juniors (n = 13) | 163 | 103 (63.19) | 60 (36.81) |
| # Hypotheses by experienced (n = 3) | 36 | 26 (72.22) | 10 (27.78) |
| **Aggregated cognitive event counts by different groups** |  |  |  |
| Total events by all | 1216 | 840 (69.08) | 376 (30.92) |
| Total events by juniors | 970 | 664 (68.45) | 306 (31.55) |
| Total events by juniors- Control (C) | 450 | 315 (70) | 135 (30) |
| Total events by juniors- VIADS (V) | 520 | 349 (67.12) | 171 (32.89) |
| Total events by experienced | 246 | 176 (71.54) | 70 (28.46) |
| **Average cognitive events per participant per hypothesis** |  |  |  |
| Mean events-junior C/hypothesis (SD) | 7.38 (5.02) | 7.68 (5.21) | 6.75 (4.66) |
| Mean events-junior V/hypothesis (SD) | 4.48 (2.43)<sup>#</sup> | 4.59 (2.69) | 4.28 (1.84) |
| Mean events-experienced/hypothesis (SD) | 6.15 (3.03)<sup>*</sup> | 5.87 (3.03) | 7 (3.06) |

Note: SD, standard deviation; <sup>#</sup> p < 0.001 between junior C and junior V; <sup>\*</sup>p < 0.01 between junior V and experienced.

### Cognitive events comparison between VIADS and non-VIADS participants

Furthermore, we compared the percentages of cognitive event count between the VIADS and non-VIADS groups among inexperienced clinical researchers (Figure 3). “Use analysis results” (31.3% vs.27.1%, p < 0.001), “Seek connections” (25.4% vs. 17.8%, p < 0.001), and “Analyze data” (22.1% vs. 21.1%) were the events with the highest percentages. The “Seek connections”, “Use analysis results”, and “Pause/think” (3.8% vs. 9.3%, p < 0.05) all show statistical differences between the VIADS and control groups by t tests. Our results indicate that the participants in the VIADS group registered higher event counts during “Preparation”, when “Analyzing results”, and when “Seeking connections”. Conversely, the control group exhibited greater event counts in categories such as “Needing further study”, “Inferring”, “Pausing”, “Using checklists”, and “Using PICOT”.

**Figure 3.**
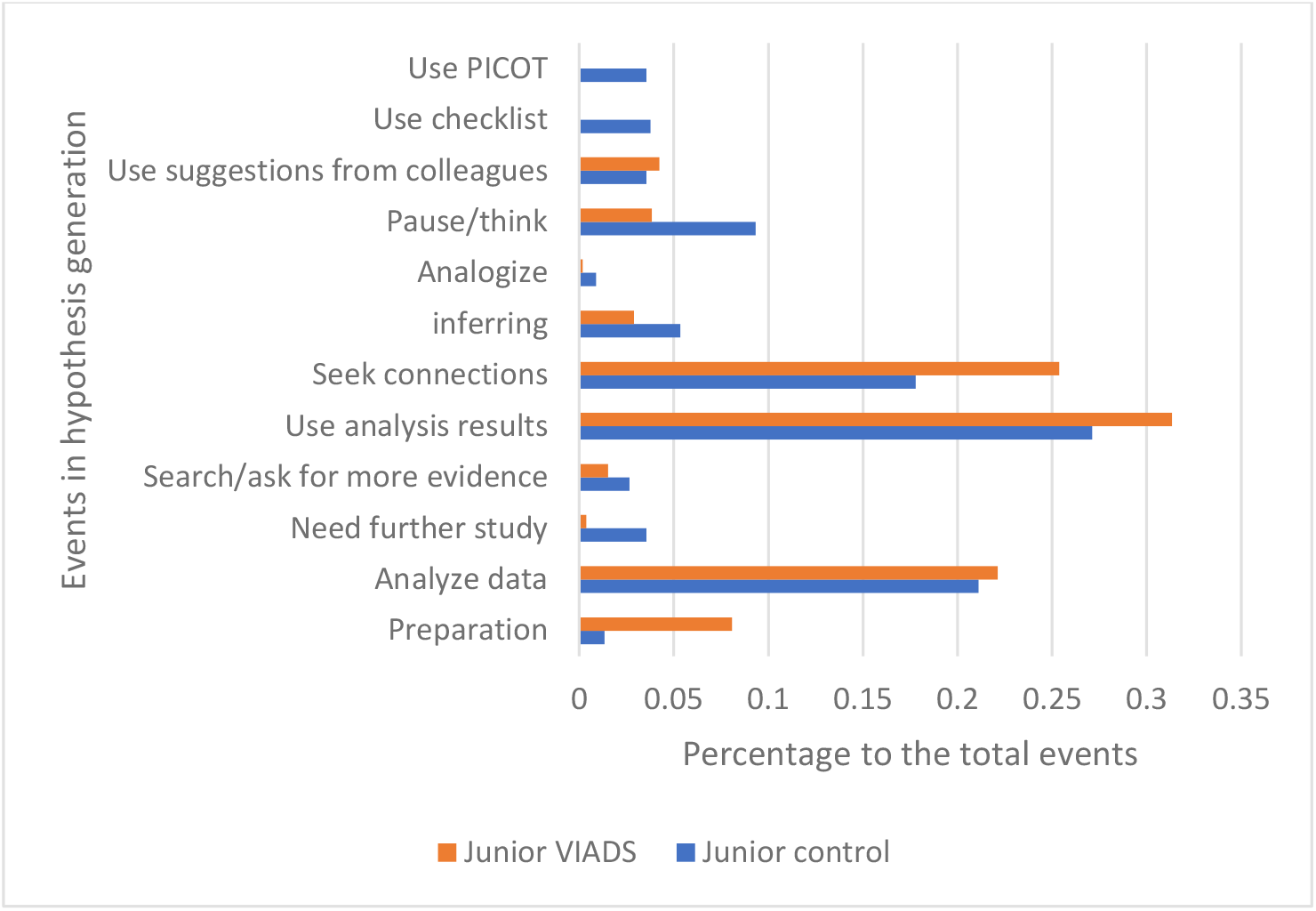
Comparing cognitive events generated by VIADS and control groups among inexperienced clinical researchers while generating hypotheses

### Cognitive events comparison between experienced and inexperienced clinical researchers

We also examined the differences between experienced and inexperienced clinical researchers regarding the percentages of cognitive events they used (Figure 4). “Use analysis results” (31.7% vs. 29.4%, p < 0.01), “Seek connections” (27.6% vs. 21.9%, p < 0.01), and “Analyze data” (17.5% vs. 21.6%, p< 0.01)) were events with the highest percentages of use. The data suggest that experienced clinical researchers exhibit higher percentages regarding these cognitive events: “Using analysis results”, “Seeking connections”, “Inferring”, and “Pausing”. Conversely, inexperienced clinical researchers demonstrated elevated percentages in cognitive events such as “Preparation”, “Data analysis”, “Utilizing suggestions”, “Utilizing checklists”, and “Utilizing PICOT”.

**Figure 4.**
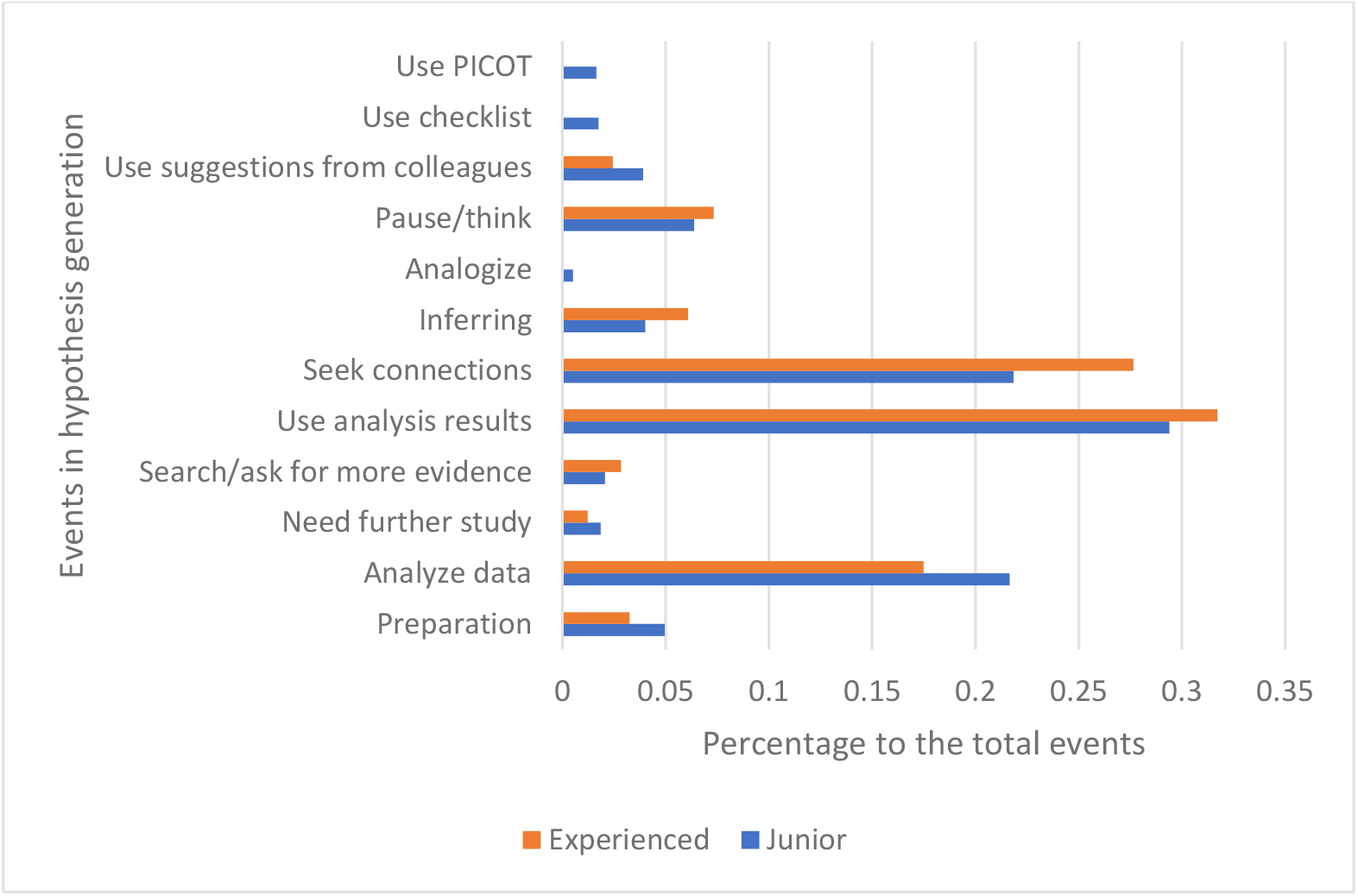
Comparison of cognitive events between experienced and inexperienced clinical researchers while generating hypotheses

### Summary of results

The inexperienced clinical researchers in the VIADS group used the fewest cognitive events to generate each hypothesis on average versus the control group (p < 0.001) and the experienced clinical researchers (p < 0.01, Table 2). The most frequently used cognitive events were “Use analysis results” (29.85%), “Seek connections” (23.03%), and “Analyze data” (20.81%) during hypothesis generation (Figure 2). It seems the inexperienced clinical researchers in the VIADS group demonstrated a similar trend to experienced clinical researchers (Figures 3 and 4).

## Discussion

### Results interpretation

Several findings of this study were notable. The experienced clinical researchers had a 10% higher percentage of valid hypotheses than the inexperienced clinical researchers (72.22% vs. 63.19%; Table 1), consistent with proposition and experience. Another interesting phenomenon is regarding the average cognitive events used by the different groups: the junior VIADS group used far fewer events per hypothesis than the control or experienced groups (4.38 vs. 7.38 vs. 6.15, Table 1) and exhibited the lowest SD. This is highly significant as it indicates that the VIADS group, despite comprising inexperienced clinical researchers, used fewer cognitive events to generate each hypothesis on average. This result supports our hypothesis that VIADS facilitates hypothesis generation. In addition, this result supports our findings that the VIADS group used a shorter time to generate each hypothesis on average [23].

Our results show clinical researchers spent ≥ 70% of cognitive events to ***<u>process evidence</u>*** during hypothesis generation. The top three cognitive events used by clinical researchers during hypothesis generation included “Using analysis results” (29.85%), “Seeking connections” (23.03%), and “Analyzing data” (20.81%, Figure 2).

Figure 3 presents the cognitive events and their distributions between the VIADS and control groups comprising the inexperienced clinical researchers. The participants in the VIADS group showed a higher number of cognitive events for interpreting the results, and the participants in the control group showed a higher number of cognitive events for external help, such as checklists and PICOT, during hypothesis generation. Figures 3 and 4 show that the VIADS group exhibits similar cognitive event trends with those of the experienced group in terms of “Using analysis results” and “Seeking connections”:

- Using analysis results:
  - VIADS versus control: 31.35% versus 27.11% (p< 0.001);
  - experienced versus inexperienced: 31.71% versus 29.38% (p < 0.01)
- Seeking connection:
  - VIADS versus control: 25.38% versus 17.78% (p< 0.001);
  - experienced versus inexperienced: 27.64% versus 21.86% (p< 0.01).

The results indicate that VIADS may help inexperienced clinical researchers move in a direction that aligns more with that of experienced clinical researchers. A more carefully designed study is needed to support or deny such a statement. However, it appears that the current quantitative evidence of cognitive events and their distributions among all cognitive events support such a trend.

### Significance of the work

We consider this study to have the following significance: 1) developed the cognitive framework for hypothesis generation in the clinical research context and provided quantitative evidence through cognitive events for the framework; 2) identified and elaborated evidence-based cognitive mechanisms that might be underneath hypothesis generation; 3) identified that experienced clinical researchers possess a considerably higher valid rate of hypothesis generated in a 2-hour window than the inexperienced clinical researchers; 4) demonstrated that VIADS may help inexperienced clinical researchers to use fewer cognitive events than participants without using in hypothesis generation, which indicates VIADS provides a structured way of thinking during hypothesis generation; and 5) established the baseline measures of cognitive events in hypothesis generation and the following events were used in descending order: processing evidence, seeking evidence, and preparation.

### Comparing to other studies

Patel et al. have explored medical reasoning through diagnoses, which have significantly influenced the design of the current study [7,8,20,22]. From their studies, we know that there were differences in the reasoning processes and thinking steps between experienced and inexperienced clinicians in medical diagnosis [9,19,22,43,44]. Therefore, we separated the participants into experienced and inexperienced groups before assigning them randomly into VIADS or control groups. The findings of this study mostly align with those of Patel et al. despite our different settings, medical diagnosis versus scientific hypothesis generation in clinical research. The experienced participants used fewer cognitive events than inexperienced participants on average, although the VIADS group used the lowest number of cognitive events despite comprising inexperienced clinical researchers.

Klahr and Dunbar’s landmark study published in 1988 [6] also enlightened our study [6]. Their study taught participants to use an electronic device. The participants had to figure out an unencountered function of the device. The process was employed to study hypothesis generation, reasoning, and testing iteratively. They concluded that searching memory and using results from prior experiments are critical for hypothesis generation. The primary differences between our studies lay in two folds: (1) the tasks for the participants (2) and the types of hypotheses generated. In the Klahr and Dunbar’s study, hypotheses had correct answers, i.e., problem-solving with one or multiple correct answers. Most likely, the participants used convergent thinking [7]. Their study used a simulated lab environment to assess scientific thinking. Conversely, the hypothesis generation in our study is open discovery without correct answers. The participants in our study used more divergent thinking during the process [7]. The hypothesis generation process in our study was substantially messier, unpredictable, and challenging to consistently evaluate comparing to their well-defined problems.

### Limitations and challenges

One of the main limitations is only three experienced clinical researchers participated in our study who generated 36 hypotheses. We compared the inexperienced and experienced groups regarding all the hypotheses and cognitive events used. However, we could not compare the cognitive events between the VIADS and control groups among the experienced clinical researchers. We made similar efforts to recruit inexperienced and experienced clinical researchers via comparable platforms; however, the recruitment results were considerably worse in the experienced group.

Another limitation of the study was that the information could be captured via the think-aloud protocol. We acknowledge that we only captured the verbalized events during the study sessions, which is a subset of the conscious process and a small portion of the real process. Our coding, aggregation, and analysis are based on the captured events.

In addition, we also faced challenges in terms of unexpected technical failure and unpredictability because this was a human-participation study. The audio recordings of two participants were partial because of a technical failure. One mitigation strategy that could be used was to conduct a test recording *each time* for every participant, which can be particularly critical if a new device is used in the middle of the study.

### Future work

Several avenues for future research emerge from our study. First, we aim to explore the sequence pattern of cognitive events to furnish additional insights into hypothesis generation. Furthermore, juxtaposing the frequencies of cognitive events with the quality evaluation results of the generated hypotheses might illuminate the potential patterns, further enriching our understanding of the process. Finally, a larger scale study encompassing a larger participant sample size and situated in a more natural environment can enhance the robustness of our findings.

## Conclusion

Experienced clinical researchers exhibit a higher valid hypothesis rate than inexperienced clinical researchers. The VIADS group of inexperienced clinical researchers used the fewest cognitive events with the lowest standard deviation to generate each hypothesis compared with experienced and inexperienced clinical researchers not using VIADS. This efficiency is further underscored by the VIADS group taking the least average time to generate a hypothesis. Notably, the VIADS inexperienced cohort mirrored the trend observed in experienced clinical researchers in terms of cognitive event distribution. Such findings indicate that VIADS may provide structured guidance during hypothesis generation. Further studies, ideally on a grander scale and in a more natural environment, could offer a deeper understanding of the process. Our research provides foundational metrics on cognitive event measures during hypothesis generation in clinical research, demonstrating the viability of executing such experiments in a simulated setting and unraveling the intricacies of the hypothesis generation process through these experiments.

## Supporting information

on the same datasets (Appendix A),

with the same study scripts (Appendix B)

initial codes and code groups (Appendix C)

initial coding principles (Appendix D)

Appendix E illustrates the processes and events

Appendix F presents individual cognitive events

## Data Availability

All data produced in the present study are available upon reasonable request to the authors

## Acknowledgments

We want to thank all participants sincerely for their precious time, courage, and expertise in helping us understand this critical but less-known hypothesis generation process better. This project received support from the National Library of Medicine (R15LM012941) and was funded partially by the National Institute of General Medical Sciences of the National Institutes of Health (P20 GM121342). The intellectual environment and research training resources provided by the NIH/NLM T15 SC BIDS4Health (T15LM013977) enriched this work.

## Appendices

Appendix A: Datasets used by participants during study sessions

Appendix B: Study session scripts followed by all participants

Appendix C: Codes and code group used during study session transcription analysis

Appendix D: Rationale and guidelines for coding data-driven hypothesis generation recordings

Appendix E: Cognitive events and their percentages during hypothesis generation in clinical research

Appendix F: Cognitive events used while generating data-driven hypotheses

