## Supplementary material for "How do clinical researchers generate data-driven scientific hypotheses? Cognitive events using think-aloud protocol": initial codes and code groups (Appendix C)

| Code | Comment | Code Group 1 | Code Group 2 | Code Group 3 | Code Group 4 | Code Group 5 | Code Group 6 | Code Group 7 | Code Group 8 | Code Group 9 | Code Group 10 | Code Group 11 | Code Group 12 | Code Group 13 |
|---|---|---|---|---|---|---|---|---|---|---|---|---|---|---|
| Analogy | | | | | | | | | | | | PROCESS_USE_EVIDENCE | | |
| Analyze evidence | | | COMPILE_EVIDENCE | | | | | | | | | | | |
| Analyze data | Include mathematic calculation, math comparison, conduct statistical tests, of think about the analytic strategies. The data can be numbers, can be texts too. | | | | | | | | | | | | | SEEK_EVIDENCE |
| Search/ask for more evidence | Currrently is doing so or need to do so | | | | | | | | | | | | | SEEK_EVIDENCE |
| Backward reasoning: from a hypothesis to evidence | | BACKWARD_REASONING | | | | | | | | | | | | |
| Building on prior hypothesis | | | | | | | INITIATE_IDEA | | | | | | | |
| Compare evidence | | | COMPILE_EVIDENCE | | | | | | | | | | | |
| Considering clarity | | | | FINALIZE_IDEA | | | | | | | | | | |
| Considering creativity/novelty | | | | FINALIZE_IDEA | | | | | | | | | | |
| Considering ethicality | | | | FINALIZE_IDEA | | | | | | | | | | |
| Considering feasibility | | | | FINALIZE_IDEA | | | | | | | | | | |
| Considering testability | | | | FINALIZE_IDEA | | | | | | | | | | |
| Consulting/conversation | | | | | FORMULATE_IDEA | | | | | | | | REFINE_IDEA | |
| Draw conclusion | | | COMPILE_EVIDENCE | | | | | | | | | | | |
| Forward reasoning: from evidence to hypothesis | | | | | | FORWARD_REASONING | | | | | | | | |
| Fresh start | | | | | | | INITIATE_IDEA | | | | | | | |
| From selected focus | | | | | | | | | | OTHER_REASONING | | | | |
| Inference | | | | | | | | | | | | PROCESS_USE_EVIDENCE | | |
| Need further study | | | | | | | | | | | | | | SEEK_EVIDENCE |
| Not obtain evidence | | | | | | | | NOT_OBTAIN_EVIDENCE | | | | | | |
| Obtain evidence | | | | | | | | | OBTAIN_EVIDENCE | | | | | |
| Pause/think | Including think | | | | | | | | | | | PROCESS_USE_EVIDENCE | | |
| Preparation | | | | | | | | | | | PREPARATION | | | |
| Seek for connection | Including to build networks conceptually | | | | | | | | | | | PROCESS_USE_EVIDENCE | | |
| Synthesize evidence | | | COMPILE_EVIDENCE | | | | | | | | | | | |
| Use analysis results | | | | | | | | | | | | PROCESS_USE_EVIDENCE | | |
| Use checklist provided in the script | | | | | | | | | | | | PROCESS_USE_EVIDENCE | | |
| Use existing file with high to low rank | Including any type of current data | | | | FORMULATE_IDEA | | | | | | | | REFINE_IDEA | |
| Use find evidence | | | | | | | | | | | | PROCESS_USE_EVIDENCE | | |
| Use PICOT framework | | | | | | | | | | | | PROCESS_USE_EVIDENCE | | |
| Use prior experience to generate idea | including prior knowledge | | | | FORMULATE_IDEA | | | | | | | | REFINE_IDEA | |
| Use suggestion from colleague | | | | | | | | | | | | PROCESS_USE_EVIDENCE | | |
| Refinement | | | | | | | | | | | | | | |
