## Supplementary material for "How do clinical researchers generate data-driven scientific hypotheses? Cognitive events using think-aloud protocol": initial coding principles (Appendix D)

Atlas.ti Code Reasoning

1. For now, only use boxes to the left of red line on the example sheet
2. Reasoning for each code
    a. Preparation- Anytime the participant is preparing the data for analysis. (pulling up the data set/VIADS chart)
        i. For VIADS group- if participant is rapidly changing the graph without looking into the data (ex: changing P-value)
        ii. For VIADS group- when participant is looking at results preview before looking at data
    b. Seek for Evidence
        i. Analyze Data- Whenever looking at data set(s) and determining things from it, BEFORE the participant generates the hypothesis/has evidence
            1. Observations
            2. Scrolling without talking
            3. Using statistical methods such as excel, generating graph, etc
            4. Asking for ICD code(s)
            5. Comparing 2 years or different codes in one year
            6. Relating to their own experience
        ii. Need Further Study- If it needs more studying to determine, and is mentioned by the researcher that they need further study.
            1. Usually one sentence long
            2. "If I had more data I would …" or "I would search PubMed" or "This would have to be investigated/researched further"

iii. Search/ask for more evidence- Asking for more evidence to help determine a hypothesis or support a claim, or physically looking for more evidence. This evidence is outside what is given in the study.

1. Participant uses website to search
2. Participant asks for evidence OUTSIDE of the data set
3. Participant wants to know more information (ex: population size)

c. Obtain Evidence/ Not obtain Evidence

d. Process/Use Evidence

i. Use Analysis Results- Using the analyzed data to make a hypothesis/observation.

1. GENERATE HYPOTHESIS

ii. Use Find Evidence- rarely used

1. Participant looks for something specific to support idea

iii. Seek for Connections- looking at connections between data points. Not as specific as inferences, but still drawing a connection between important information.

1. Participant is speculating, listing many reasons, “maybe …”
2. Participant relating to experience
3. Comparing or connecting 2 things

iv. Inferencing- Making educated guesses from data/observations. Must be concrete inferences.

1. “One could assume …”

v. Analogy- Drawing parallels between one similar thing (i.e. personal research) to another similar thing in the data set.

1. Participant finds similar (NOT same) pattern with 2 codes
2. Relate to an experience or observation

vi. Pause/think- When the participant pauses (stops talking or is not doing anything on screen)

1. Participant is silent

vii. Use suggestions from colleagues (Dr. Jing)- Using suggestions from Dr. Jing

1. Conversation between participant and Dr. Jing

viii. Use Checklist- Using the checklist provided at the end of the study script.

ix. Use PICOT- Using the PICOT method to make hypotheses.

1. Always used with use analysis results code
