## Appendix E illustrates the processes and events for "How do clinical researchers generate data-driven scientific hypotheses? Cognitive events using think-aloud protocol"

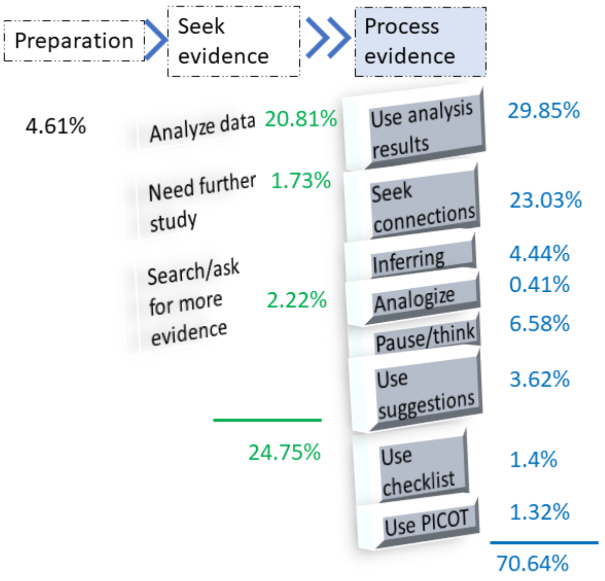


**Appendix E** Cognitive events and their percentages during hypothesis generation in clinical research (based on coding)
