## Appendix F presents individual cognitive events for "How do clinical researchers generate data-driven scientific hypotheses? Cognitive events using think-aloud protocol"

**Appendix F** Cognitive events used while generating data-driven hypotheses

| | All hypotheses | | Valid only | |
|---|---|---|---|---|
| **Cognitive events** | **# of events** | **% of events** | **# of events** | **% of events** |
| **Preparation** | 56 | 4.61 | 8 | 1.1 |
| **Seek evidence** | 301 | 24.75 | 169 | 23.25 |
| **Analyze data** | 253 | 20.81 | 138 | 18.98 |
| **Need further study** | 21 | 1.73 | 10 | 1.38 |
| **Ask for more evidence** | 27 | 2.22 | 21 | 2.89 |
| **Process evidence** | 859 | 70.64 | 550 | 75.65 |
| **Use analysis results** | 363 | 29.85 | 231 | 31.77 |
| **Seek connections** | 280 | 23.03 | 178 | 24.48 |
| **Infer** | 54 | 4.44 | 44 | 6.05 |
| **Analogize** | 5 | 0.41 | 4 | 0.55 |
| **Pause/think** | 80 | 6.58 | 46 | 6.33 |
| **Use suggestions** | 44 | 3.62 | 20 | 2.75 |
| **Use checklist** | 17 | 1.4 | 12 | 1.65 |
| **Use PICOT** | 16 | 1.32 | 15 | 2.06 |
| **Total cognitive events** | 1216 | | 727 | |

Note: green indicates an increased percentage of cognitive events in valid versus all hypotheses, and orange represents a decrease; PICOT: patient, intervention, comparison, outcome, type of study.
